# Promoter methylation of LDLR is not a common cause of familial hypercholesterolemia

**DOI:** 10.64898/2026.11.16.26368244

**Authors:** Gabrielle N. Altman, Alejandro Martin-Trujillo, Luke B. Hesson, Damon A. Bell, Amanda J. Hooper, Andrew J. Sharp

## Abstract

**Background/Objectives:** Familial hypercholesterolemia (FH) is a widely prevalent inherited disorder defined by chronically high LDL-C concentrations and a significantly increased risk of cardiovascular disease. While FH is primarily an autosomal dominant disease caused by pathogenic mutations in *LDLR, APOB*, or *PCSK9*, approximately 20–60% of clinically diagnosed patients are mutation-negative. A previous survey of over 23,000 methylation profiles identified a differentially methylated region in the promoter of *LDLR*. This study aimed to investigate whether constitutional hypermethylation of the *LDLR* promoter is a recurrent cause of FH in patients lacking identifiable pathogenic variants.

**Methods:** We screened 308 mutation-negative clinically confirmed FH patients for promoter hypermethylation in *LDLR*. We used Combined Bisulfite Restriction Analysis (COBRA) with genomic DNA from peripheral blood samples. The assay assessed methylation at *TaqI* sites within a 332 bp region of the *LDLR* promoter.

**Results:** No instances of *LDLR* promoter hypermethylation were detected among the 308 mutation-negative FH patients. Statistical analysis yielded an upper bound of 1.19% (95% Clopper–Pearson CI) for the prevalence of this specific epimutation in our cohort.

**Conclusions:** Our results indicate that *LDLR* promoter hypermethylation at this specific locus is not a common cause of the FH phenotype in mutation-negative patients. These findings imply that while methylation in this region may be rare or not significantly contribute to the disease phenotype, the diagnostic gap in FH may be explained by other factors, such as methylation in other regions, polygenic risk, structural variants, or non-canonical regulatory mutations.

## 1. Introduction

Familial hypercholesterolemia (FH) is an inherited disease with elevated serum low-density lipoprotein (LDL) cholesterol levels, which increase the risk of premature atherosclerotic cardiovascular disease [1]. It affects approximately 1 in every 300 individuals worldwide [2]. Clinical diagnosis of FH is often based on the Dutch Lipid Clinic Network (DLCN) criteria [3]. This point-based scoring system incorporates family history, clinical history, LDL-C levels, and physical examination findings to classify individuals as unlikely FH (score <3), possible (3–5), probable (6–8), or definite FH (>8). FH is primarily an autosomal dominant disorder with multiple known causative pathogenic variants in the genes *LDLR, APOB*, and *PCSK9* [4]. Autosomal recessive mutations in *LDLRAP1* are an extremely rare cause of FH [4]. FH is most commonly caused by pathogenic variants in *LDLR*, which encodes the low-density lipoprotein receptor (LDLR), a key receptor that regulates blood LDL-C levels [1,5]. Pathogenic variants in *LDLR* that cause FH typically lead to loss or reduction of receptor function, resulting in decreased receptor-mediated LDL uptake and thus the high LDL levels seen in FH [1,5]. Despite well-established genetic testing, known pathogenic variants in FH-associated genes are detected in approximately 40–80% of patients with definite FH (DLCN > 8) and in fewer patients with probable FH. This diagnostic gap suggests that alternative variants or mutational mechanisms underlie the disease in a subset of cases [5–8]. Identifying the underlying molecular cause in the remaining mutation-negative patients is critical for enabling accurate cascade testing of family members and refining cardiovascular risk assessment.

One relatively underexplored mechanism in relation to FH is promoter hypermethylation. DNA hypermethylation (i.e., increased methylation) occurring in gene promoters is usually associated with transcriptional silencing, and several studies have demonstrated that constitutional epimutations can underlie some human diseases [9,10]. For example, constitutional hypermethylation in the promoter of *MLH1* and transcriptional silencing of the affected allele occur in a small proportion of selected cohorts of patients with Lynch syndrome who are negative for pathogenic coding variants [11,12].

Despite evidence that epigenetic variation contributes to human disease, the prevalence and distribution of rare epigenetic variation in the human population are understudied. To address this, a previous study conducted a survey of methylation profiles in over 23,000 individuals, representing the largest cohort of methylomes at the time [13]. The survey identified differentially methylated regions (DMRs) with outlier methylation profiles generated using Illumina 450k HumanMethylation BeadChip (450k array) for >23,000 genomes. This analysis identified four samples (prevalence of ∼1 in 6,000) with a hypermethylated DMR spanning >200 bp in the promoter of *LDLR* comprising seven consecutive array probes, validated by bisulfite sequencing. As promoter hypermethylation often leads to transcriptional silencing of genes, *LDLR* promoter hypermethylation could potentially explain some undiagnosed cases of FH.

DNA methylation has been proposed as an additional mechanism contributing to the FH phenotype, particularly in patients without detectable pathogenic variants in canonical FH genes. For example, promoter methylation of *ABCA1* has been associated with HDL-C levels and coronary artery disease in FH, supporting a role for epigenetic regulation in lipid metabolism [14]. A handful of other studies have also begun to explore the role of epivariation in FH, examining additional functional regions in FH-related genes [15,16] or investigating the methylome genome-wide [17]. While these studies highlight the importance of characterizing the role of epivariation in FH, they were often limited by small cohort sizes or focused on regions outside the *LDLR* promoter region of interest here.

Consequently, it remains uncertain whether constitutional *LDLR* epimutations represent a substantial cause of FH. In the present study, we addressed this by analyzing a larger cohort of FH patients without known pathogenic variants and focusing on a single, well-defined region of the *LDLR* promoter. We hypothesized that inactivating promoter hypermethylation in *LDLR* explains a subset of mutation-negative FH cases. To test this, we screened for *LDLR* promoter hypermethylation in a large cohort of 308 mutation-negative FH patients.

## 2. Results

An analysis of 308 DNA samples from mutation-negative FH patients was conducted using the COBRA assay. In all 308 of these samples, we observed gel electrophoresis bands around 330 bp, which is the length of the undigested PCR product, consistent with an absence of methylation at the restriction sites (Figure 1). Therefore, we observed zero cases with methylation (0/308) with an exact two-sided 95% Clopper– Pearson confidence interval for the proportion of FH patients with this epimutation of 0– 1.19% (Table 1).

**Table 1.** Combined bisulfite restriction analysis (COBRA) results of *LDLR* promoter methylation in FH patients without known pathogenic variants.

|  | **N** | **N with methylation** | **% methylated (95% CI)** |
|---|---|---|---|
| FH cases, mutation-negative | 308 | 0 | 0 (0 – 1.19) |

**Figure 1.**
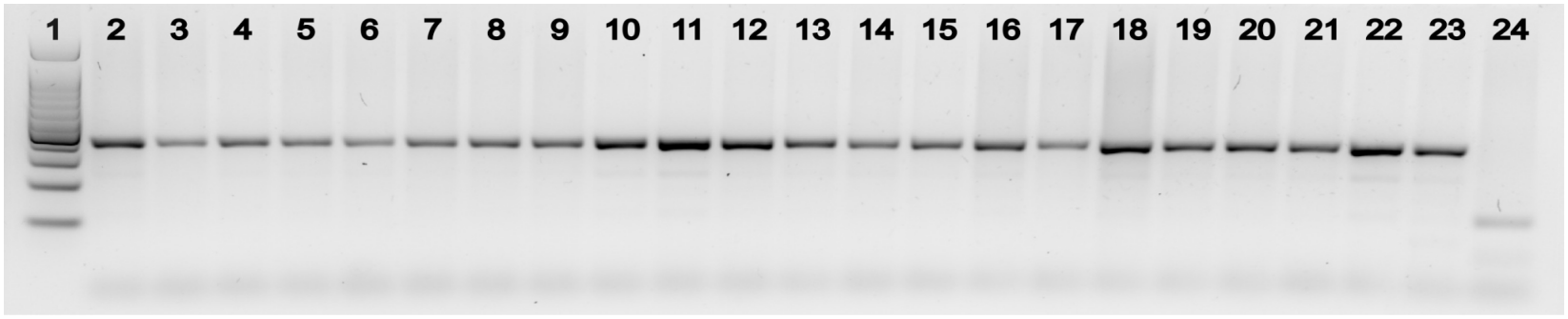
Representative COBRA gel for *LDLR* promoter methylation. Lane 1, 100 bp DNA ladder; lanes 2–23, bisulfite-converted PCR products from mutation-negative FH patients after restriction enzyme digestion; lane 24, bisulfite-converted PCR products from methylated positive control after restriction enzyme digestion. Patient samples show a single undigested band, consistent with the absence of methylation at the restriction sites. In contrast, the positive control displays the characteristic smaller digestion fragments, consistent with the presence of methylation at the restriction sites.

## 3. Discussion

Based on the prevalence of FH in the population (∼1 in 300) and the previously observed prevalence of the *LDLR* promoter hypermethylation in unselected cohorts (∼1 in 6,000), a fully penetrant causal relationship would predict that approximately 5% of FH cases carry this epimutation. Given that our cohort only contains mutation-negative FH cases, which are estimated to represent ∼20 – 60% of all clinically diagnosed FH cases, we would expect this epimutation to account for 8.3 – 25% of the individuals in our cohort. Among 308 mutation-negative FH patients, this would correspond to ∼25 – 77 expected samples with this epimutation; however, we did not identify any cases with the epimutation. Use of a two-sided Clopper–Pearson 95% confidence interval places an upper bound of 1.19% on the prevalence of this epimutation in our cohort, well below the expected prevalence if the methylation were a common, highly penetrant, causal mechanism for FH. Even under a conservative assumption that this epimutation would be present in 8.3% of mutation-negative FH samples, the probability of observing zero positives in 308 samples is 3✕10-12 (one-sided exact binomial test). Taken together, these data suggest that this epimutation is unlikely to be a frequent cause of FH in mutation-negative patients.

However, there are several technical and biological reasons why these results should not be taken as proof that *LDLR* promoter methylation is completely absent or unimportant in FH. First, COBRA is less sensitive than several other methylation detection methods. Because our assay relies on methylation at specific *TaqI* restriction sites, it has limited utility if only a subset of CpGs within the recognition motifs are methylated or if the hypermethylation exists in a mosaic state. In those scenarios, the resulting digestion fragments may be too small, faint, or sparse to be reliably visualized on an agarose gel. Additionally, if methylation occurs within this amplicon region, but not encompassing the *TaqI* sites themselves, it would not be detected by this assay. While this region of interest overlaps six of the seven Illumina 450K array probes that detected hypermethylation in the previous methylome survey study, only one of these probes overlaps with a *TaqI* recognition motif. CpG sites within a DMR are typically co-methylated, with neighboring CpGs showing correlated methylation states and co-methylation decreasing with distance [18]. In our previous work, bisulfite sequencing of a subset of this *LDLR* promoter region in a hypermethylated sample showed co-methylation of CpG sites not covered by the 450k array probes or our COBRA assay [13]. However, single-CpG–resolution studies and epiallele analyses have also demonstrated substantial heterogeneity within such regions, where adjacent or contiguous CpG sites can display discordant methylation states or complex epiallelic patterns despite falling within the same DMR [19]. Thus, our COBRA assay provides a relatively coarse, site-specific readout of methylation rather than assessing all CpGs across the entire DMR. We therefore cannot exclude the possibility that more nuanced methylation patterns exist within this promoter segment that were not captured by our restriction-based approach.

One limitation of our analysis is that we focused only on a single, relatively small region of the *LDLR* promoter and did not assess methylation in the remainder of the *LDLR* promoter, other *LDLR* regulatory elements, or in other FH-related genes. A few other studies have explored additional regions of *LDLR*, other genes, or methylation genome-wide. For example, promoter methylation of *ABCA1* was associated with HDL-C levels and coronary artery disease in FH [14], and a targeted study of 61 samples found higher methylation in a second *LDLR* CpG island among mutation-negative patients compared to controls [15]. Another investigation of 131 FH patients and 23 controls found higher methylation compared to controls at selected sites in exons of *LDLR, PCSK9*, and *LDLRAP1* and associations with cardiovascular events, though it found no major differences between mutation-positive and mutation-negative FH patients [16]. Notably, these targeted studies did not assess the specific promoter region investigated in the present work. Lastly, one study assessed methylation in 78 mutation-negative and 58 *LDLR*-mutation-positive FH patients genome-wide, rather than using a targeted approach [17]. They found one significantly hypomethylated CpG site in *CPT1A* in mutation-negative FH patients, but no significant methylation changes in canonical lipid genes, including *LDLR*, compared with mutation-positive patients. However, this study was limited by a relatively small sample size and the absence of FH-negative control samples, reducing power. Additionally, the large epivariation screen conducted by our group that initially identified this *LDLR* promoter region as a candidate DMR also detected a single instance of promoter hypermethylation in *APOB* (1 out of ∼23,000 individuals) [13]. These findings, together with previous studies, demonstrate the need to investigate epimutations outside of this specific promoter region.

In addition, our study was performed on DNA extracted from peripheral blood, which may not fully reflect methylation patterns in other tissues more directly relevant to *LDLR* expression and LDL-C metabolism. Tissue-specific methylation, age-related epigenetic drift, and differences in blood cell composition could all influence the detectability of promoter methylation in blood [20,21].

Beyond promoter hypermethylation, several alternative mechanisms may contribute to the phenotype in mutation-negative cases across all canonical FH genes. This includes complex structural variants that are not detected by standard diagnostic sequencing done in FH patients, as well as variants in distal regulatory elements. Many unexplained cases may be associated with variants of uncertain significance (VUS) that currently lack sufficient data for pathogenic classification. Alternatively, some mutation-negative FH cases may be polygenic rather than monogenic in origin. Several studies have demonstrated that a high polygenic load of common LDL-raising variants is associated with a clinical diagnosis of FH [22–24]. Future research characterizing these alternative genetic and regulatory drivers, as well as investigating novel candidate genes, remains essential for closing the diagnostic gap in FH.

To our knowledge, the present work represents the largest targeted epivariation screen focused on mutation-negative FH patients to date. Taken together, these data suggest recurrent *LDLR* promoter hypermethylation at this specific locus does not appear to be a common, highly penetrant cause of FH in our cohort, although rarer, subtler, or differently localized methylation changes in this region cannot be excluded, given the limitations of our assay. Larger multi-tissue studies using more sensitive methylation technologies will be required to fully characterize the spectrum and clinical impact of epivariation in FH.

## 4. Materials and Methods

### Patients and samples

Peripheral blood samples from 308 consented patients with clinically diagnosed FH with Dutch Lipid Clinic Network Criteria Scores (Median 6, IQR 4-8) who did not have a likely pathogenic or pathogenic variant identified were collected from the BioBank for Inherited Disorders of Lipid Metabolism at Royal Perth Hospital in Western Australia. Genomic DNA was isolated from peripheral blood leukocytes using the Wizard Genomic DNA Purification Kit (Promega, Madison, WI, USA). Routine FH genetic testing, including Sanger sequencing of *LDLR, APOB*, and *PCSK9*, and multiplex ligation-dependent probe amplification of *LDLR*, did not identify any pathogenic variants. This study was reviewed and approved by the IRB of the Icahn School of Medicine at Mount Sinai (Study ID: STUDY-22-01301).

### Combined Bisulfite Restriction Analysis (COBRA)

A combined bisulfite restriction analysis (COBRA) [25] assay was developed to determine the presence of methylation in a defined region of the *LDLR* promoter in an economical, high-throughput, and readily interpretable way. The target region is a 332 bp segment of the *LDLR* promoter, containing four *TaqI* recognition sites and 15 CpG sites, overlapping six of the seven Illumina 450K array probes that detected hypermethylation in the previous methylome survey study (**Figure 2**). One of the *TaqI* recognition sites directly overlaps a CpG assayed by the 450k array. Primers were designed using the bisulfite-converted sequence of the region without any overlapping CpG dinucleotides in the primer binding sites, so that the region is amplified irrespective of methylation status. The primer sequences used were 5’-GAGAAATTTTAGGAGGATTTTTTAGAAGATG-3’ (forward primer) and 5’-ACCTACTATATCCTAACTAAAAACCCTA-3’ (reverse primer).

**Figure 2.**
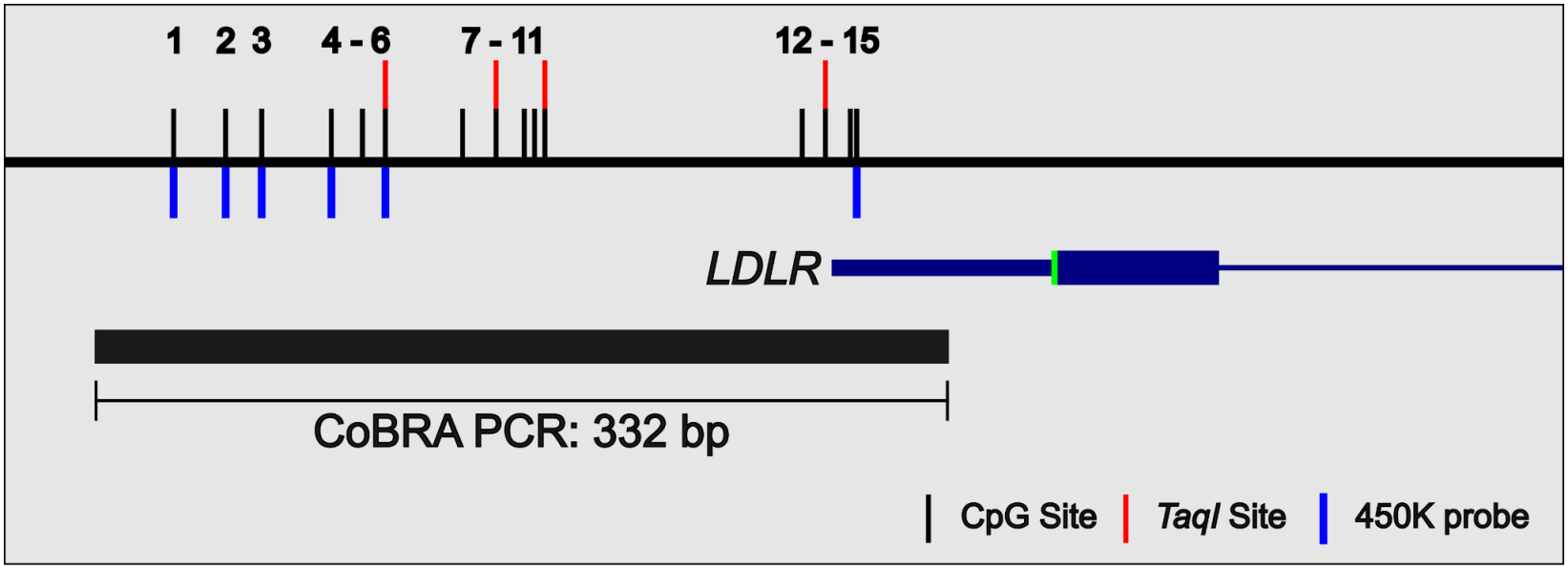
COBRA assay targets the *LDLR* promoter. Schematic of the region of interest spanning the LDLR promoter. The 332 bp COBRA PCR amplicon is shown as a black bar. The amplicon overlaps 15 CpG sites (black tick marks) and 4 *TaqI* restriction sites, which overlap CpGs and are indicated by red caps on the corresponding CpG markers. Six of these CpG sites are represented by Illumina 450K probes (blue tick marks). Structure of the *LDLR* gene is shown by blue horizontal bars of varying thickness, corresponding to the 5’ UTR, exon 1, and intron 1 (left to right).

For each sample, 1 µg of DNA was subjected to sodium bisulfite conversion using the EZ-96 DNA Methylation Kit, Deep Well (Zymo Research) according to the manufacturer’s protocol, with a final elution volume of 12 µL. Bisulfite-converted DNA (2 µL) was PCR-amplified using the designed primers. PCR reactions (25 µL) contained HotStarTaq DNA Polymerase (QIAGEN), 10x PCR buffer, 10 µM of each dNTP, and 10 µM of each primer. Reactions were cycled using 95 °C for 10 min; 45 cycles of 95 °C for 30 s, 56 °C for 30 s, and 72 °C for 30 s; followed by a final extension at 72 °C for 10 min. The resulting 332 bp PCR product was digested with the TaqI restriction enzyme (ThermoFisher) in a 30 µL reaction containing 10X TaqI Buffer and 1 U TaqI and incubated at 65 °C for 2 hours. Sodium bisulfite treatment converts unmethylated cytosines to uracils, while methylated cytosines remain unconverted. TaqI recognizes the sequence TCGA and is therefore sensitive to bisulfite-induced changes of the overlapping CpG. After bisulfite conversion, unmethylated TaqI sites become TTGA and are no longer recognized by the enzyme, while methylated TaqI sites retain the recognition sequence TCGA. Therefore, the enzyme digests the PCR products only at methylated recognition sequences. Digested products were resolved on a 2.5% agarose gel with a 100bp DNA ladder (Promega) and visualized with a GelDoc Go Imaging System (Bio-Rad). Any sample with evidence of potential methylation was repeated using a doubled PCR reaction volume, then subjected to the digestion protocol both with and without restriction enzyme, and the paired reactions were run side by side on the same gel. All reactions and gels were run with one well/lane containing the products of the COBRA assay run on CpG methylated human DNA (ThermoFisher) as a positive control, which is enzymatically methylated with CpG Methyltransferase (M.SssI), which methylates all CpG sites.

If all four TaqI sites in the 332bp region are methylated, the digestion of the PCR product generates fragments of approximately 114, 109, 47, 43, and 19 bp, with the 109 and 114 bp fragments easily visualized on a 2.5% agarose gel (Figure 1). Samples with methylation on one allele will show both the undigested 332 bp band and the smaller digestion fragments, whereas samples lacking methylation at these CpG sites in the TaqI recognition motifs show only the undigested 332 bp product.

## Data Availability

All data produced in the present study are available upon reasonable request to the authors.

## Author Contributions

Conceptualization, A.J.S and L.B.H.; methodology, L.B.H., A.J.S., and A.M.T.; investigation, G.N.A; resources, D.A.B and A.J.H; formal analysis, G.N.A.; writing—original draft preparation, G.N.A; writing—review and editing, G.N.A., A.M.T, L.B.H, D.A.B., A.J.H, and A.J.S.; visualization, G.N.A; supervision, A.M.T. and A.J.S.; project administration, A.J.S.; funding acquisition, A.J.S. All authors have read and agreed to the published version of the manuscript.

## Institutional Review Board Statement

The study was conducted in accordance with the Declaration of Helsinki and approved by the Institutional Review Board of the Icahn School of Medicine at Mount Sinai (STUDY-22-01301; October 24^th^, 2022).

## Informed Consent Statement

Informed consent was obtained from all subjects involved in the study.

## Data Availability Statement

The raw data supporting the conclusions of this article will be made available by the authors upon reasonable request.

## Abbreviations

The following abbreviations are used in this manuscript:

FH: Familial hypercholesterolemia
LDL-C: Low-density lipoprotein
VUS: Variants of uncertain significance
DLCN: Dutch Lipid Clinic Network
DMR: Differentially methylated region
*LDLR*: Low-density lipoprotein receptor
COBRA: Combined bisulfite restriction analysis
450k array: Illumina 450k Human Methylation BeadChip Bp Base pair
CI: Confidence interval
PCR: Polymerase chain reaction

